# Strict Lower Bound on the COVID-19 Fatality Rate in Overwhelmed Healthcare Systems

**DOI:** 10.1101/2020.04.22.20076026

**Authors:** Bruce A. Bassett

**Affiliations:** African Institute for Mathematical Sciences, Muizenburg, Cape Town, 7950, South Africa; Department of Maths and Applied Maths, University of Cape Town, Rondebosch, Cape Town, 7700; South African Radio Astronomical Observatory, Observatory, Cape Town, 7295; South African Astronomical Observatory, Observatory, Cape Town, 7295

## Abstract

The Infection Fatality Rate (IFR) for COVID-19 is a poorly known, yet crucial, aspect of the disease. Counting only current deaths in a region and assuming everyone in that region is infected provides an absolute lower bound on the IFR. Using this estimator for New York City, Lombardy and Madrid yields strong bounds on the average IFR in overwhelmed health systems. Their combined 35, 152 deaths implies an IFR ≥ 0.14% averaged over 25.1 million people. This is the best-case scenario and conclusively demonstrates that COVID-19 is more deadly than influenza. The actual value of the average COVID-19 IFR is likely to be higher than this bound.

## 1 Lower Bound on IFR

The Infection Fatality Rate (IFR) - *the probability of dying given infection* - for COVID-19 is one the most important questions facing humanity in early 2020, implicitly driving policies around social intervention strategy that materially affect the economic and physical health of hundreds of millions around the world.

Yet estimating the true value of the IFR during the pandemic is complicated by multiple factors. The strong age gradient and comorbidity dependence of the disease makes generalisation from small groups of infected people, such as on the *Diamond Princess*, difficult (Russell T. W., 2020). Conversely the true number of infections is currently poorly understood in much larger and more representative groups. Case Fatality Rates (CFR) - *the probability of death having tested positive* - for COVID-19 vary strongly with the number of tests per million undertaken. Figure (1)^1^ shows how the crude CFR^2^ for countries decreases with increasing number of tests per million population.

In general IFR ≤ CFR with the two coinciding when everyone in a country is tested. Since this is infeasible, finding lower bounds on the IFR allows us to squeeze the true range of the IFR.

**Figure 1:**
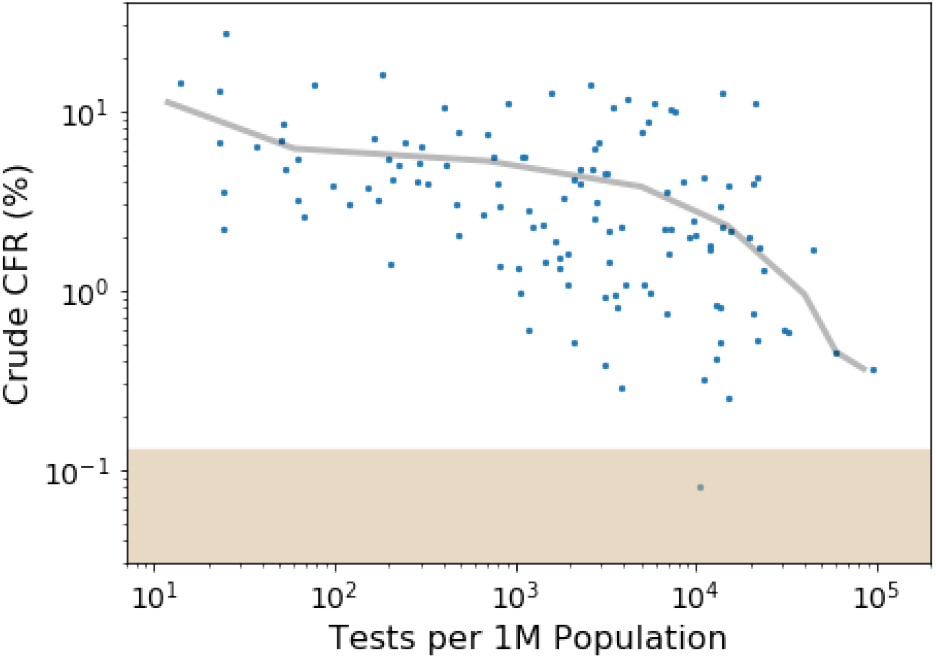
Crude Case Fatality Rate (cCFR) for countries vs number of tests per million population; the line shows the mean cCFR in bins. The IFR region excluded by the joint analysis of Lombardy, Madrid and New York City (IFR < 0.13), is shown in beige. cCFR underestimates the true CFR since it does not include future deaths stemming from current infections.

A strict lower bound on the IFR can be obtained for any region X by:

1. Counting only confirmed, or highly-probable COVID-19 deaths in X
2. Assuming everyone in X has been infected

Assuming everyone is infected, and not including any likely future deaths, gives the lowest possible estimate of the IFR. The actual IFR will always be larger than the ratio of these two factors since infecting the entire population is almost impossible and the number of deaths is monotonically increasing ^3^.

Applying this to most regions yields lower limits that are far too small to be interesting: as an extreme example, applying it to the whole world yields the bound IFR *>* 0.002%. Even Hubei, which was the first place hard hit, only tells us that IFR > 0.008%. Neither of these lower limits is particularly useful. However, COVID-19 infections have now progressed in severity to such an extent in several regions that this method yields useful lower bounds on the IFR, as we now discuss.

## 2 Regional Analysis

Lombardy, Madrid and New York City have all experienced significant deaths due to COVID-19. Across these three regions there were 35, 152 deaths by 22 April 2020, leading to an average lower limit of IFR ≥ 0.14%, computed over a combined population of 25.1 million people. This represents the best-case-scenario for COVID-19 and is a strict demonstration that the average COVID-19 IFR for large, heterogeneous groups is significantly higher than that of influenza. For example, estimates of the 2017 influenza IFR are around 0.002% ^4^ while the fatality rate for symptomatic cases in the 2009 H1N1pdm09 pandemic was estimated to be about 0.1%, with the IFR smaller still (Wong, 2013).

Assuming that everyone in a region has been infected is useful since there can be no missing infections in the absence of travel. However, a more realistic upper estimate of the number of infections is provided by the final predicted fraction likely to be infected, which for *R*_0_ = 2.4 was estimated to be approximately 81% of the population for the UK and USA (Ferguson N., 2020). Using this *predicted* fraction increases the average lower bound on the IFR to 0.17% over all three regions. However, unlike our earlier estimate, this number is now model-dependent since it depends on the value of *R*_0_ and assumes the validity of models/simulations, and hence could be wrong. Table (1) shows all our results combined.

**Table 1:**
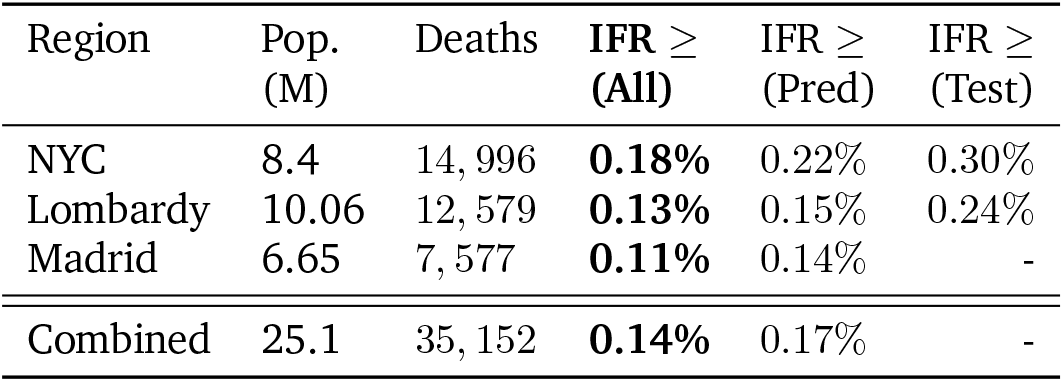
Estimated IFR lower bounds (in %) by region and assumed fraction of population infected using data as of 22 April 2020. “All” assumes everyone is infected and provides the most robust lower bound, or put another way, the best possible case scenario. “Pred” corresponds to 81% of the full population while “Test” uses 60% (NYC) and 51% (Lombardy) of the population as estimates of infected fractions respectively. No testing data is available for Madrid.

| Region | Pop. (M) | Deaths | $IFR \geq$ (All) | $IFR \geq$ (Pred) | $IFR \geq$ (Test) |
| --- | --- | --- | --- | --- | --- |
| NYC | 8.4 | 14,996 | **0.18%** | 0.22% | 0.30% |
| Lombardy | 10.06 | 12,579 | **0.13%** | 0.15% | 0.24% |
| Madrid | 6.65 | 7,577 | **0.11%** | 0.14% | - |
| Combined | 25.1 | 35,152 | **0.14%** | 0.17% | - |

We now consider each region individually.

### 2.1 New York City

As of 22 April 2020, New York City had recorded 14, 996 COVID-19 deaths, of which about two thirds are confirmed through positive COVID-19 tests, and one third list COVID-19 as cause of death^5^ from a population of approximately 8.4M. This yields IFR lower bounds of 0.12% using only confirmed COVID-19 deaths and 0.18% if both confirmed and probable COVID-19 deaths are included^6^. Using the theoretically predicted infected fraction of 81% increases the lower bound to IFR ≥ 0.22%.

Detailed testing results for New York State are available for each county every day^7^. Since no county has had more than 60% positive tests on any day (Bronx reached that level on 30 March 2020), and since most testing has been undertaken on more serious, symptomatic patients, we can get a tighter bound on the IFR by taking 60% as our estimate of the maximum number of residents who have been infected at some point.

This leads to the tightest bound of IFR ≥ 0.30%. However, this does not account for false negative results which are significant for PCR (Wikramaratna P., 2020). It is therefore possible that the actual prevalence of COVID-19 could be higher than the observed maximum testing fraction. This is therefore the least robust lower bound on the IFR.

### 2.2 Madrid

As of 22 April 2020, Madrid had recorded 7, 577deaths in a population of approximately 6.65M, leading to lower bounds of IFR *≥* 0.11% and 0.14% under the asssumptions that all and 81% of the population are infected respectively.

### 2.3 Lombardy

As of 22 April 2020, Lombardy had recorded 12, 579 deaths in a population of approximately 10.06M. Assuming that 100% and 81% of the population are infected gives IFR lower bounds of 0.13% and 0.15% respectively. Further, testing results show that the maximum positive fraction of tests for the region was 50.9% (which occurred on 20 March 2020)^8^. Using the latter as an estimate of the upper bound on the total infected population leads to an IFR lower bound of 0.24%. Again this result is subject to concerns about false negatives and is less rigorous and trustworthy^9^, though likely to actually be closer to the true IFR for COVID-19.

## 3 Discussion

We have argued that the Infection Fatality Rate (IFR) of COVID-19 in overwhelmed medical systems must be at least 0.14% on average by combining data from Lombardy, Madrid and New York City, establishing that COVID-19 is more deadly than influenza. The estimate is maximally conservative: it counts only confirmed and probable deaths and assumes that everyone in the regions included in the analysis is infected. The estimate is also robust as it comes from a sample of over 25 million people in three different countries, with a range of age pyramids and comorbid disease distributions.

This estimate is conservative since it ignores future deaths and the high likelihood that significant portions of the populations in the regions are likely not infected. As a result, this lower bound will steadily increase as some of the current and future infected populations die. Based on the relatively slow decrease in daily deaths in countries such as China, the final IFR bound from this method is likely to be significantly larger than 0.14%.

One potential limitation of our analysis is that all three regions included have experienced some level of overwhelm of their healthcare systems, manifesting as shortages of some combination of beds, oxygen, ventilators, medicines and health care workers, leading to an increase in the computed IFR. How large of an effect this is is currently unknown. While the IFR for patients in healthcare systems that are not overwhelmed is very interesting, it is arguably more useful for governments to know the IFR in the overwhelmed case, since this is the likely scenario they will face should they decide to significantly relax social interventions such as lockdown.

An important assumption in this work has been that travel to and from the affected regions has not significantly affected death tolls, i.e. that there has not been an in/outflux of travelers who later died. This could potentially happen if, e.g., the regions offered to help other affected regions by taking on large numbers of severely ill patients. However, the converse - infected people leaving to go to be in areas with less overwhelmed medical systems - seems more likely. As a result the lower bounds presented here seem fairly robust to this effect.

Finally it is interesting to note that our lower bounds from the three worst-hit regions in Italy, Spain and the USA, namely (0.18%,0.11%,0.13%), are fairly similar. If the true population-averaged IFR is much higher, e.g. 1.5%, this would be a somewhat surprising co-incidence. One potential explanation could be that a similar fraction of the population is currently infected in each region but given that the epidemics started at different times this would be surprising unless the spread has been shut off by herd-immunity or some other effect. Such a causal explanation would also require that the fraction of missing deaths to be similar in all regions. If this is the case we expect the true IFR to be closer to the numbers in the “Pred” column of Table (1).

## Data Availability

All data used are publicly available from third-party sources.

## Acknowledgements

I thank Mike Berkman for discussions which lead to the main idea of this paper and Andrew Bateman, Linda Camara, Fernando Camilo, Sue Cleary, Lucy Jamieson, Ben Holder, David Kaiser, Joe Okleberry and Jon Shock for useful discussions or comments. This paper was created using Latex with an adapted version of the Wenneker article template^10^.

## Footnotes

1 Data on 9 April 2020; http://www.worldometers.info/coronavirus/

2 Crude CFR is defined to be the number of deaths divided by the number of confirmed cases at the same time *t*, i.e. does not account for the delay between infection and death.

3 Barring a major reassignment of deaths.

4 https://www.cdc.gov/nchs/fastats/flu.htm, https://www.cdc.gov/nchs/data/nvsr/nvsr68/nvsr68_09-508.pdf

5 https://www1.nyc.gov/site/doh/covid/covid-19-data.page

6 A further 10,023 deaths are of unknown origin. A fraction of these may have been indirectly caused by COVID-19 due to lack of access to medical facilities (perceived or real) and add to the indrect IFR from COVID-19.

7 https://covid19tracker.health.ny.gov/views/NYS-COVID19-Tracker/NYSDOHCOVID-19Tracker-Map?

8 https://github.com/pcm-dpc/COVID-19, https://datastudio.google.com/u/0/reporting/91350339-2c97-49b5-92b8-965996530f00/page/RdlHB, https://ourworldindata.org/covid-testing#italy

9 Since people recover from COVID-19, some unknown fraction of PCR tests will return negative for people whose viral load has dropped too much to be detected. As a result it is likely possible to construct scenarios where the true fraction of the population who have been infected is higher than the fraction in any tests.

10 Available from http://www.LaTeXTemplates.com

## Notes

### Competing Interest Statement

The authors have declared no competing interest.

### Funding Statement

No external funding has been received to support this research. The authors have not received payment of services from any third party for any aspect of the submitted work.

